# Detection of carbapenem resistance genes in *Campylobacter coli* and *Campylobacter jejuni* isolated from chickens, and diarrheic children aged less than five years in Kampala city, Uganda

**DOI:** 10.1101/2023.09.10.23295341

**Authors:** Walter Okello, Ann Nanteza, Felix Opiyo, Justin Okello, Lesley Rose Ninsiima, Peter Marin, David Onafruo, Patrick Pithua, Clovice Kankya, Terence Odoch

**Affiliations:** Department of Biosecurity Ecosystems and Veterinary Public Health, College of Veterinary Medicine, Animal Resources and Biosecurity (COVAB), Makerere University, P.O. Box 7062, Kampala, Uganda; International Institute of Tropical Agriculture, P.O. Box 7878, Kampala, Uganda; Department of Population Health Sciences, Virginia-Maryland College of Veterinary Medicine, Virginia Polytechnic Institute and State University, Blacksburg, 24061, United States

**Keywords:** Carbapenem resistance genes, *Campylobacter coli*, *Campylobacter jejuni*, Chickens, Diarrheic children aged less than five years

## Abstract

*Campylobacter* species are recognized as one of the significant causative agents of global foodborne illnesses and potential reservoirs for dissemination of antimicrobial resistance due to their zoonotic nature. Unlike other bacteria such as *Klebsiella pneumoniae, E. coli, Enterobacter*, etc., *Campylobacter* has shown limited or absent resistance to Carbapenems, critically important “last resort” antibiotics. This distinct resistance profile prompted this investigation into the prevalence of Carbapenem resistance genes in *Campylobacter* species, specifically *Campylobacter coli* (*C. coli*) and *Campylobacter jejuni* (*C. jejuni*).

Analyses were conducted on 292 archived *C. coli* and *C. jejuni* isolates obtained from chickens and diarrheic children under five years of age in Kampala city, Uganda. The primary objectives included assessment of phenotypic susceptibility of the isolates to Meropenem and Imipenem using the Kirby Bauer disk diffusion method, as well as determination of the occurrence of four selected Carbapenem resistance genes (blaVIM, blaNDM-1, blaIMP, and blaOXA-48) using multiplex polymerase chain reaction (PCR).

Interestingly, despite the observed phenotypic susceptibility to Meropenem and Imipenem in all the *Campylobacter* isolates, 29.8% harbored at least one of the four selected Carbapenem resistance genes, including blaVIM, blaNDM-1, blaIMP, and blaOXA-48. The prevalence of the resistance genes was 55(28.1%) in chickens and 15(38.5%) in children. Notably, blaVIM was the predominant gene, detected in 57.1% of the isolates, followed by blaNDM-1 (11.4%), blaIMP (8.6%), and blaOXA-48 (5.7%). Coexistence of multiple resistance genes was also observed, with blaVIM and blaIMP present in 10.0% of the isolates, and blaVIM and blaNDM-1 in 5.7%. One isolate displayed simultaneous presence of blaNDM-1, blaVIM, and blaIMP.

This study uncovered a previously unexplored realm in *Campylobacter* research, identifying Carbapenem resistance genes in *Campylobacter* in Uganda. The identification of these resistance genes, despite the apparent phenotypic susceptibility to Carbapenems, signifies the presence of a substantial reservoir of carbapenem resistance genes in *Campylobacter*.

## Introduction

Antimicrobial resistance (AMR) is an increasing global threat to public health, posing a significant risk to vulnerable populations, including neonates, critically ill patients, and those with weakened immune systems [1]. The World Health Organization [2] has identified AMR as a global health security concern, emphasizing the need for action across government sectors and society as a whole. In the United States alone, the Centers for Disease Control (CDC) estimated that AMR-related direct healthcare costs could reach up to $20 billion annually, with additional costs of up to $35 billion per year in terms of lost productivity [3]. Moreover, a report released by the Uganda National Academy of Sciences in 2015 highlighted the worsening trends of resistance and the diminishing effectiveness of commonly used antibiotics in Uganda [4].

In response to the rising AMR threat, Uganda has taken significant steps to combat the spread of antibiotic resistance. Notably, the country has developed a National Antimicrobial Resistance Action Plan spanning from 2018 to 2023. This action plan outlines a comprehensive strategy to tackle AMR across multiple sectors and stakeholders, including the health sector, agriculture, animal husbandry, and environmental management. The implementation of this plan aims to curb the misuse and overuse of antibiotics, enhance surveillance and monitoring of resistant pathogens, improve infection prevention and control practices, and promote responsible antimicrobial use in both humans and animals. Even though the Uganda AMR national action plan, supports the analysis, dissemination, and sharing of surveillance data and Information on AMR to facilitate, decision-making on diagnoses and treatments in clinical public health, veterinary practice, environment, and wildlife laboratories and food technologies, no data on molecular mechanisms associated with resistance to Carbapenems in *Campylobacter* has been generated.

*Campylobacter* infections are a major cause of bacterial gastroenteritis worldwide [5], with *Campylobacter jejuni* and *Campylobacter* coli being the primary species responsible for human disease. The Centers for Disease Control and Prevention (CDC) estimated that a substantial number of infections, approximately 300,000 cases per year in the United States alone, are caused by drug-resistant *Campylobacter* strains [6]. This alarming trend underscores the urgent need for comprehensive surveillance and understanding of AMR mechanisms in *Campylobacter* especially in low-income settings such as Uganda.

Poultry, including chickens, plays a critical role in the epidemiology of AMR, especially in the context of *Campylobacter* species [7]. The transmission of resistant *Campylobacter* from poultry to humans is of significant concern, as it may compromise the effectiveness of commonly used antibiotics for the treatment of these infections. The proximity of humans to poultry and the potential for direct or indirect contact with contaminated poultry products contribute to the spread of AMR at the animals-humans-environment interface [8], amplifying the global burden of antimicrobial resistance.

The emergence and dissemination of carbapenem resistance in Gram-negative bacteria including *Campylobacter* pose an alarming threat, as carbapenems are often considered as last-line treatment options for severe bacterial infections [9].

Resistance to Carbapenems is commonly due to the production of Carbapenemases such as; veronica integron metallo-beta-lactamase types (VIM), imipenemase (IMP) types, *Klebsiella pneumoniae* Carbapenemase (KPC), oxacillinase_48 (OXA_48), and New Delhi metallo-beta-lactamase-1 (NDM-1), encoded by Carbapenem resistance determining genes (CRDGs):*blaVIM, blaIMP, blaKPC, blaOXA_48*, and *blaNDM-1*, respectively [10]. Resistance to Carbapenems may also be due to plasmid AmpC beta-lactamases in combination with ESBLs, mutations, and altered expression of porins. The presence of carbapenem resistance genes in *Campylobacter* strains raises concerns about the potential dissemination of these resistance determinants in the environment, which could further contribute to the global spread of AMR.

Therefore, this research aimed to investigate the phenotypic resistance patterns to selected carbapenems and the prevalence of CRDGs in *C. coli* and *C. jejuni* isolated from chickens and diarrheic children aged less than five years in Kampala city, Uganda. This research will provide critical insights into the dynamics of carbapenem resistance in this pathogen and contribute to informing effective strategies for its control and prevention, ultimately protecting public health.

## Materials And Methods

### Study design

This was a cross-sectional study that aimed at determining phenotypic resistance patterns against Meropenem and Imipenem and detecting four Carbapenem resistance genes (*blaOXA-48, blaNDM-1, blaVIM and blaIMP*) in archived samples of *Campylobacter species, C. coli, and C. jejuni* isolates at Food Hygiene Laboratory, Makerere University, College of Veterinary Medicine, Animal Resources and Biosecurity (COVAB).

### Study population

The study was conducted on all 292(48 from children and 244 from chickens) isolates of *Campylobacter species* previously stored at -40°C in the Food Hygiene Laboratory at the Department of Biosecurity, Ecosystems and Veterinary Public Health in 2021. The bacterial isolates had been recovered from feacal and rectal swab samples from diarrheic children aged five years and below who were attending Mulago National Referral Hospital and Kisenyi Health Centre III and from cloacal swabs of chickens in the childrens’ homes or neighborhoods in Kampala city, Uganda, between 2020 and 2021. These isolates were obtained using the method as described by [11]. Briefly, faecal samples were cultured on Exeter broth and transferred to Modified Charcoal Cefoperazone Deoxycholate Agar (MCCDA) through 0.45μm Nitrocellulose filters. Suspected *Campylobacter* colonies were sub -cultured on blood agar and Identification was based on colony morphology.

### Retrieval of *Campylobacter* species isolates from storage

The isolates and associated data were first accessed on 1^st^ September 2022. Retrieval of isolates and analysis of phenotypic antibiotic susceptibility patterns were performed from the Food Hygiene Laboratory at COVAB using the method by [11]. All the 292 cryopreserved samples of *Campylobacter species* in Microbank (Oxoid) with 15% glycerol were resuscitated in nutrient broth by pipetting 100µl of the glycerol stock in which the isolates were archived into 1ml of nutrient broth. Incubation was done at 42°C for 24 hours under microaerophilic conditions created by Campygen^®^ (Oxoid, UK) gas-generating pack in an anaerobic jar. After 24-hour incubation, 100µl of the broth culture was transferred onto a 0.45micrometer nitrocellulose membrane filter placed on sterile MCCDA (Blood-Free agar) (Oxoid, UK) containing *Campylobacter* selective supplement SR0155 (Oxoid, UK). The fluid was spread on the filter using a sterile micro loop and allowed to drain through for 30 minutes. Incubation was then done for 24 hours at 42°C under microaerophilic conditions created by Campygen^®^ (Oxoid, UK) in an anaerobic jar. A pool of colonies from the MCCDA was emulsified in nutrient broth (Oxoid, UK) and transferred to the Molecular Laboratory at COVAB for DNA extraction and identification of *Campylobacter species* using PCR.

### Determination of phenotypic antimicrobial susceptibility patterns against Meropenem and Imipenem in *Campylobacter* species isolates

All the archived isolates once resuscitated were subjected to antimicrobial susceptibility testing using the Kirby-Bauer disk diffusion susceptibility testing Method [12]. The standardised bacterial inoculum using 0.5 Mcfarland standards was grown on Mueller-Hinton agar (Oxoid, UK) in the presence of two Carbapenem antibiotics (Imipenem 10µg, Meropenem 10µg) (Oxoid, UK) and incubated for 24 hours at an ambient temperature of 42°C in a microaerophilic condition provided by Campygen^®^. At the end of incubation, the zone inhibition diameters were measured using a ruler and a pair of dividers. The results of the zone inhibition diameters were interpreted as resistant, intermediate, and susceptible following the Clinical and Laboratory Standards Institute (CLSI), 2021 guidelines [13]. Details of the Carbapenem breakpoints are summarized in Table 1.

**Table 1:**
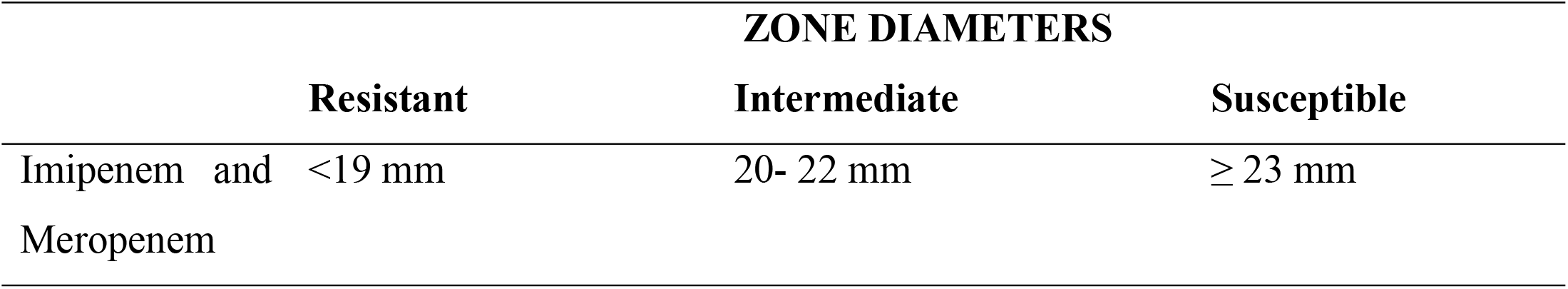
Standard zone diameters for Imipenem and Meropenem CLSI, 2021 [13].

### Molecular identification of *Campylobacter* species and detection of carbapenem resistance genes

#### *Campylobacter* DNA Extraction

Wizard genomic DNA purification kit from PROMEGA (USA) was used to extract DNA based on the manufacturer’s instructions: Centrifugation of 1ml of the overnight culture was done for two minutes at 13,000 rpm and the supernatant was discarded. Nuclei Lysis Solution (600μl) was added to the pellet and incubated for five minutes at 80°C, then cooled to room temperature. Three microliters of RNase solution were added and incubated at 37°C for 60 minutes, then cooled to room temperature (25°C). Two hundred microliters of Protein precipitation solution were added, vortexed, incubated on ice for five minutes, and centrifuged at 13,000 rpm for three minutes. The supernatant was transferred to a clean tube containing 600μl of room-temperature isopropanol, centrifuged, and the supernatant decanted. Seventy percent ethanol (600μl) was added to the pellet, mixed, and centrifuged for two minutes at 13,000 rpm. The ethanol was aspirated, and the pellet was air-dried for 10 minutes at 25°C. Finally, the DNA pellet was rehydrated in 100μl of Rehydration Solution for one hour at 65°C.

### Identification of genus *Campylobacter*, species *C. coli* and *C. jejuni* by PCR

Using a multiplex PCR, *Campylobacter specie*s 16S RNA gene, specific genes for *C. coli and C. jejuni* (*ask* and *cj0414* respectively) were amplified as described by [14]. Briefly, 1x of wifi Taq mix, 0.25µM of each forward and reverse primers (Table 2), and 1.5µl of the template DNA were mixed and topped up with PCR water to a 25µl reaction volume. The amplification was carried out in a thermocycler (Bio Rad, USA) using the following conditions: initial denaturation at 95°C for 15 minutes followed by 25 cycles of denaturation at 95°C for 30 seconds, annealing at 58°C for 1 minute, extension at 72°C for 1 minute and a final extension at 72° C for 7 minutes.

**Table 2:**
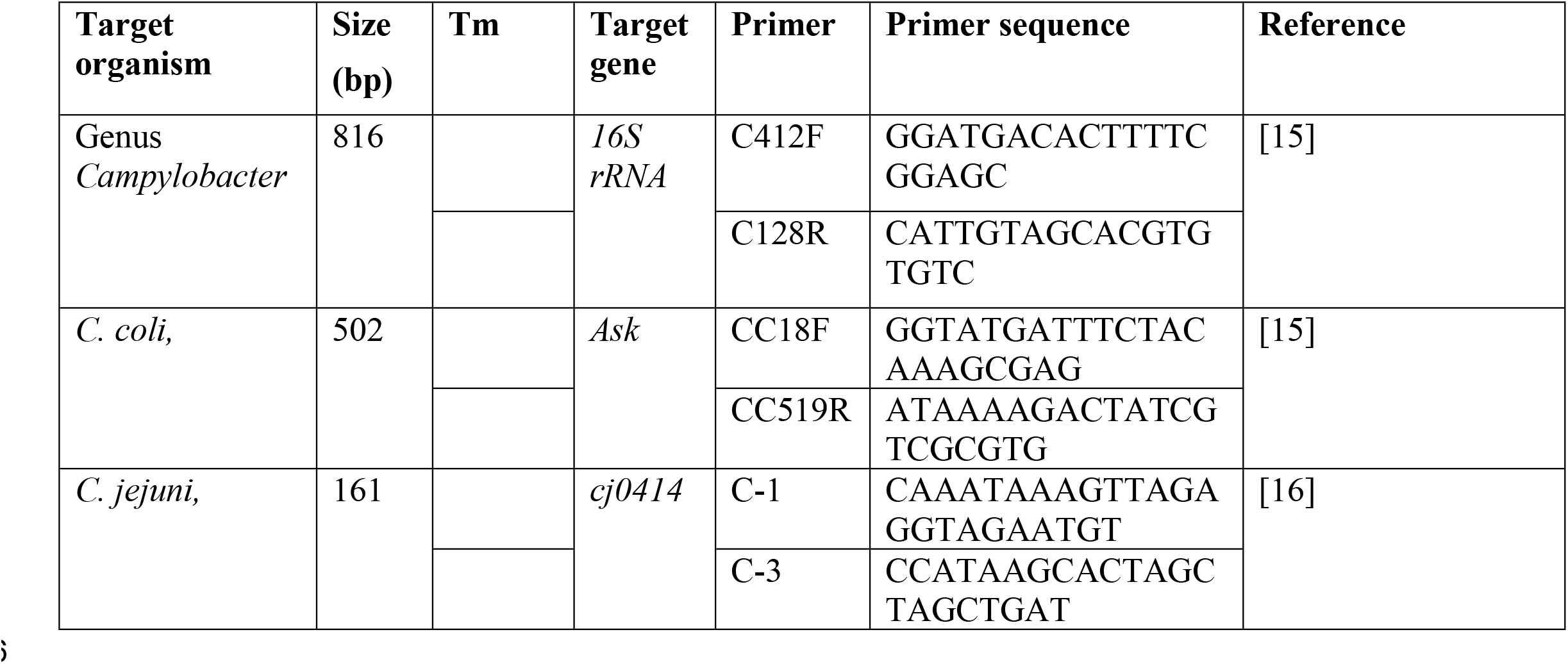
Primers used in the amplification of *Campylobacter* species, *C. coli* and *C. jejuni*.

The PCR amplicons were run on 2% agarose gel. The gel was run in Tris-acetate EDTA (TAE) buffer at 125 voltage for 35 minutes in the presence of gel red for staining the DNA. The gel was then visualised under a ultra-violet gel illuminator.

### Multiplex PCR detection of Carbapenem resistance genes

Only positive samples for *Campylobacter species* 16S *RNA, C, coli*, and *C. jejuni* were subjected to the detection of resistance genes. My Taq Mix (Bioline, Alvinston, Canada) of 2x Concentration was used to perform the Multiplex PCR to detect the four resistance genes (blaOXA*_48, blaNDM-1, blaIMP*, and blaVIM) using eight primers shown in table 3. These four genes were selected because of their clinical significance and being the most commonly reported carbapenem resistance genes in other bacteria such as the *Enterobacteriaceae*.

**Table 3:**
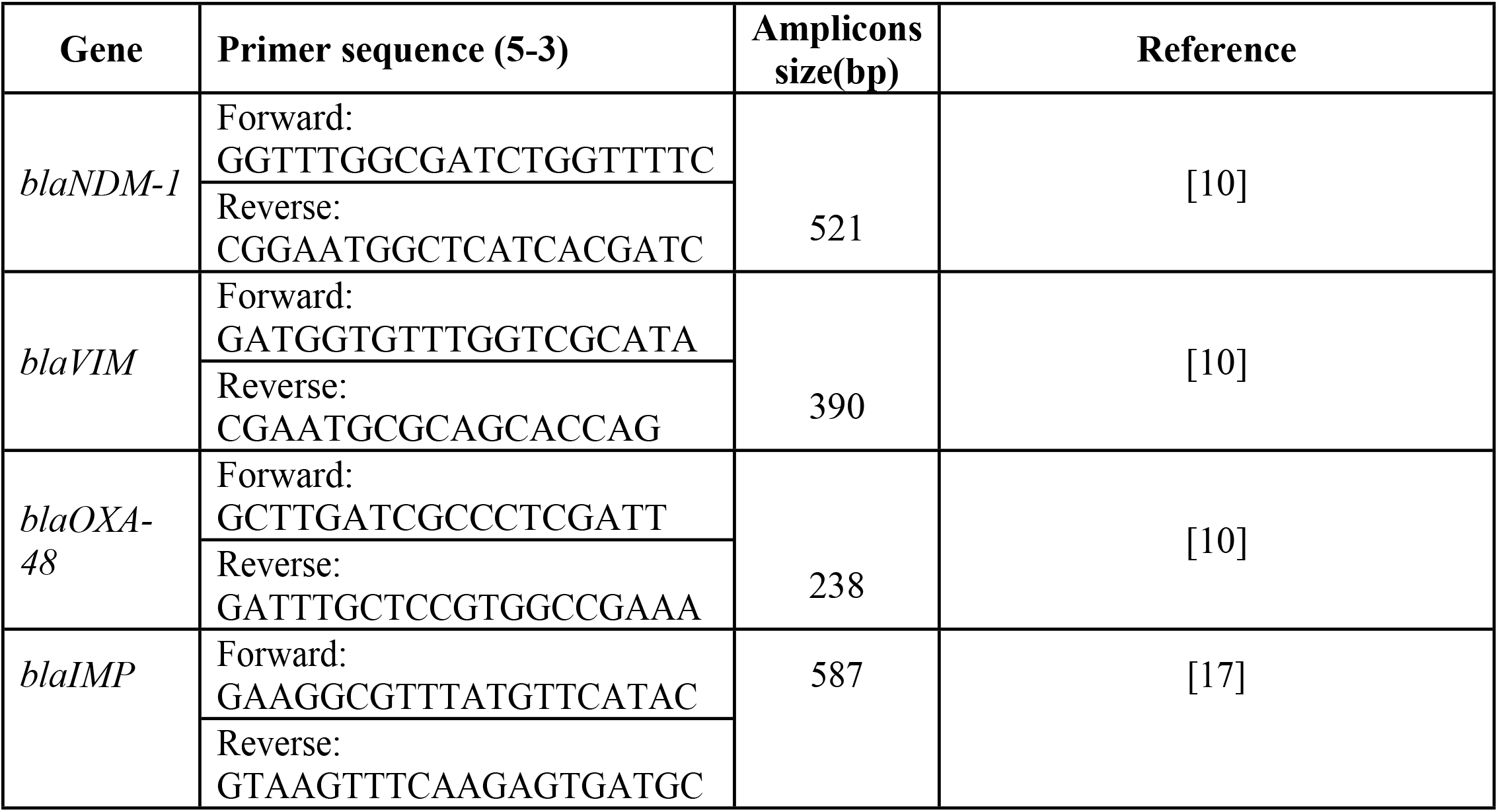
Primers for amplification of four selected Carbapenem resistance genes.

A total PCR reaction of 12.5µl was used comprising 6.25 µl of 2x My Taq Mix, 0.25 µl of 10µM of both Forward and reverse primer, and 2µl of DNA template and nuclease-free water was used to top up to 12.5 µl total reaction mix. The following PCR programs were used, initial denaturation at 94°C for 5 minutes, followed by 40 cycles of denaturation at 94°C for 45 seconds, annealing at 50°C for 1 minute, extension at 72°C for 1 minute, one final cycle of extension at 72°C for 10 minutes and hold at 4°C for infinity. Electrophoresis was done to detect the presence of the resistance genes and run on 2% agarose gel at 100V, 300MA for 50 minutes, and DNA stained with 3x Gel Red. The gel was then visualised under a UV gel illuminator.

### Data analysis

The phenotypic antibiotic susceptibility patterns and Carbapenem resistance genes for each isolate were recorded as per sample identification numbers. All results obtained were verified for any clerical errors by comparing the final lab report results with the raw data generated in the lab that detailed every step undertaken before concluding on the results reported. After the verification process, the generated data were coded and entered in Microsoft Excel and then exported to STATA 14 software for analysis. Data analysis was conducted in STATA version 14, proportion differences were computed using the Chi-square test, and statistical differences were determined at 0.05 level of significance and 95% confidence level.

### Ethical considerations

This study did not require ethical approval as it focused on archived isolates. However, permission to use the isolates was sought from the Principal Investigator of the primary study that obtained the isolates. The primary study underwent review and approval by the School of Health Sciences, Makerere University Research Ethical Committee and received approval from the School of Health Sciences Research Ethics Committee (SHSREC REF. 2020/034), also uploaded as ‘others’. The approved informed consent included the option to utilize stored isolates for further research purposes. Written consent was obtained from care takers of the children whose stool samples were collected in the primary study. All files containing electronic data were password-protected. Laboratory identification numbers previously used for sample isolation, which concealed patients’ identities, were retained during this analysis.

## Results

### Phenotypic antimicrobial resistance patterns of *Campylobacter species* against Meropenem and Imipenem

All 292(100%) isolates of *C. coli* and *C. jejuni* were phenotypically susceptible to both Meropenem and Imipenem antibiotics, (Zone of inhibition diameters ranged from 33-48mm, Mean and SD= 41.5 4.2 respectively)

### Molecular identification of *Campylobacter species, C. coli* and *C. jejuni*

Out of the 292 isolates of *Campylobacter species* included in this study, 288(98.6%) of the isolates were positive for genus *Campylobacter* 16S RNA gene, of which 31(10.8%) were *C. jejuni*, 204(70.8%) were *C. coli*, 50(17.4%) mixed isolates of *C. coli* and *C. jejuni* and 3(1.0%) were genus *Campylobacter* but neither *C. coli* nor *C. jejuni*. Of the 235 isolates confirmed as *C. coli* and *C. jejuni*, 39(16.60%) were from children aged less than five years and 196(83.40%) were from chicken. 38(97.44%) of the 39 isolates from children were *C. coli*, 1(2.56%) was *C. jejuni*; and 166(84.69%) of the 196 isolates from chicken were *C. coli* and 30 (15.31%) were *C. jejuni*. A representative agarose gel for multiplex PCR amplification of *Campylobacter* species 16S RNA genes, *C. coli*, and *C. jejuni* specific genes is shown in figure 1.

**Fig 1:**
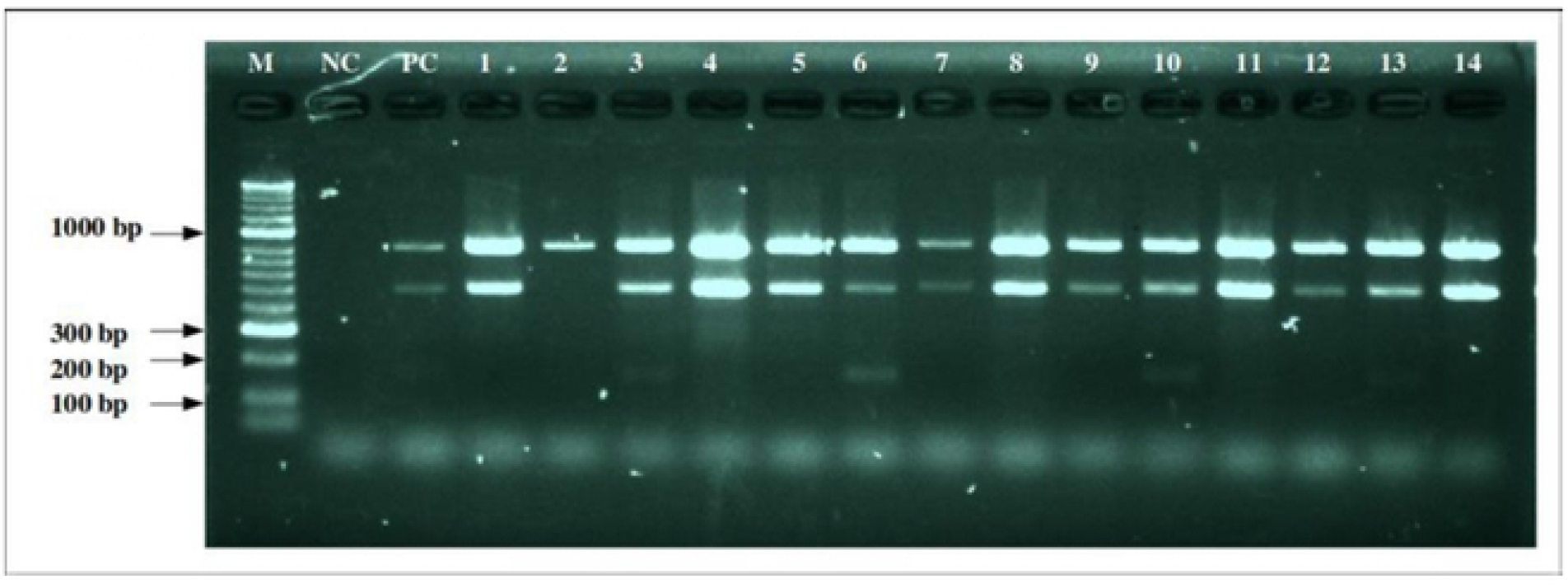
Representative agarose gel for multiplex PCR amplification of Campylobacter species 16S RNA genes, C. coli, and C. jejuni specific genes Lane M: Molecular marker-50bp (Bioline, Alvinston, Canada). Lane NC: Negative control (Distiled water). Lane PC: Positive control-C. coli (ATCC 33559, Thermo Fisher, US) positive control. Lanes 1-14 test samples. Lane 2 is Campylobacter species different from C. coli and C. jejuni; Samples 1-14 are all positive for Campylobacter species 16S RNA gene, and sample 2 is a Campylobacter species different from C. coli and C. jejuni. Lanes 3, 6, 10, and 13 have both C. coli and C. jejuni. Campylobacter species were detected on a gel by the presence of 816bp band, while C. coli and C. jejuni had bands of 502bp and 161bp, respectively.

### Prevalence of Carbapenem resistance genes in *C. coli* and *C. jejuni*

Out of the 235 isolates of *C. coli* and *C. jejuni*, 70(29.8%, 95% Conf. Interval= 0.2424904 0.3598934) were positive for at least one of the four carbapenem resistance genes targeted in this study with a higher prevalence in *C. jejuni*, 16(51.6%) than in *C. coli*, 54(26.5%). The prevalence of the resistance genes was 55(78.5%, CI= 0.6712809-0.8681365) in chickens and 15(21.4%, CI= 0.1318635-0.3287191) in children.

Overall, the most predominant gene was blaVIM 40(57.1%), followed by blaNDM-1 8(11.4%), blaIMP 6(8.6%) and blaOXA-48 was 4(5.7%). A combination of blaVIM and blaIMP was 7(10.0 %), blaVIM and blaNDM-1 4(5.7%), and 1(1.4%) had blaNDM-1, blaVIM and blaIMP. The difference in the prevalence of the genes in *C. coli* and *C*.*jejuni* was statically significant (Chi-square value= 4.0358 and P-value = 0.045) as shown in table 4 and 5. A representative agarose gel for multiplex PCR amplification of selected carbapenem resistance genes (blaVIM, blaIDM-1, blaOXA-48, and blaIMP) in *C. coli* and *C. jejuni* is shown in figure 2.

**Table 4:**
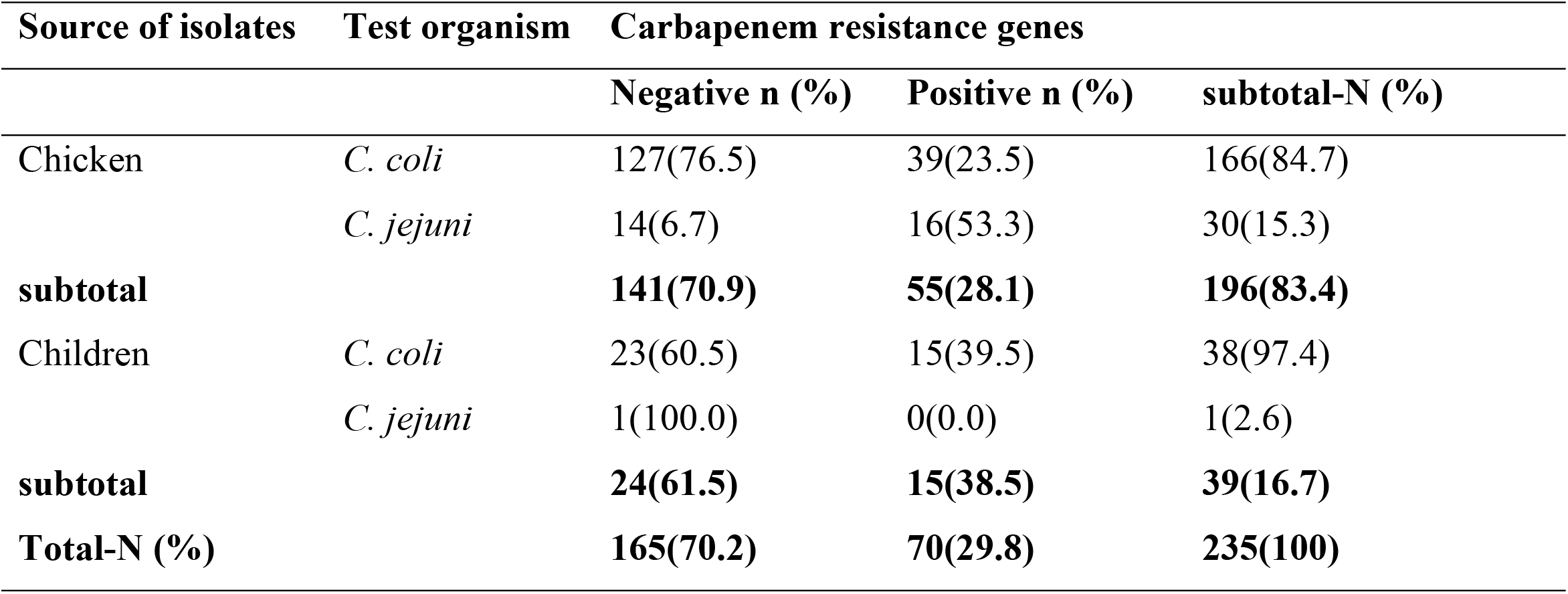
Frequency of Carbapenem resistance genes in *C. coli and C. jejuni* isolates.

| Source of isolates | Test organism | Carbapenem resistance genes |  |  |
| --- | --- | --- | --- | --- |
|  |  | Negative n (%) | Positive n (%) | subtotal-N (%) |
| Chicken | *C. coli* | 127(76.5) | 39(23.5) | 166(84.7) |
|  | *C. jejuni* | 14(6.7) | 16(53.3) | 30(15.3) |
| **subtotal** |  | **141(70.9)** | **55(28.1)** | **196(83.4)** |
| Children | *C. coli* | 23(60.5) | 15(39.5) | 38(97.4) |
|  | *C. jejuni* | 1(100.0) | 0(0.0) | 1(2.6) |
| **subtotal** |  | **24(61.5)** | **15(38.5)** | **39(16.7)** |
| **Total-N (%)** |  | **165(70.2)** | **70(29.8)** | **235(100)** |

**Table 5:**
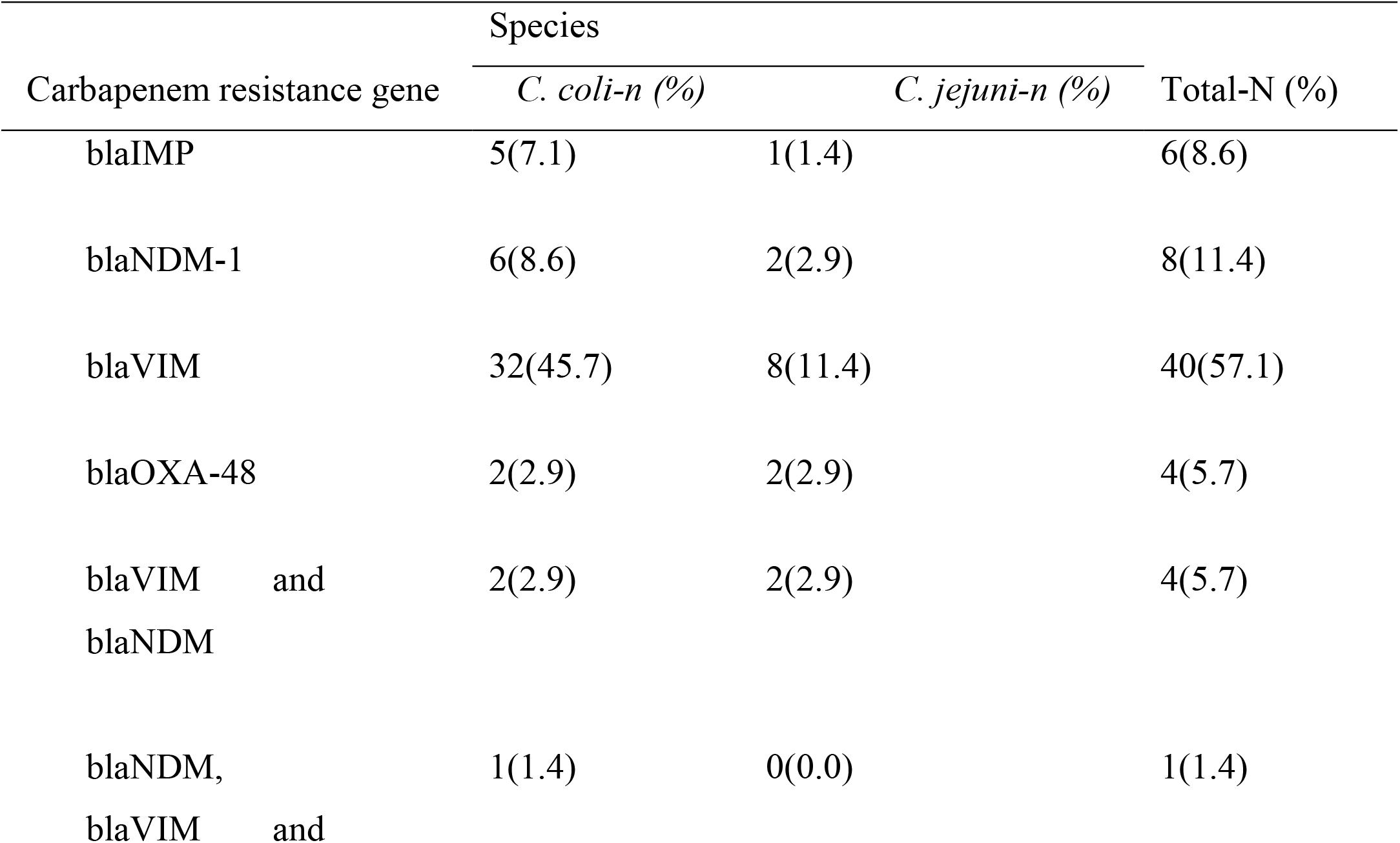
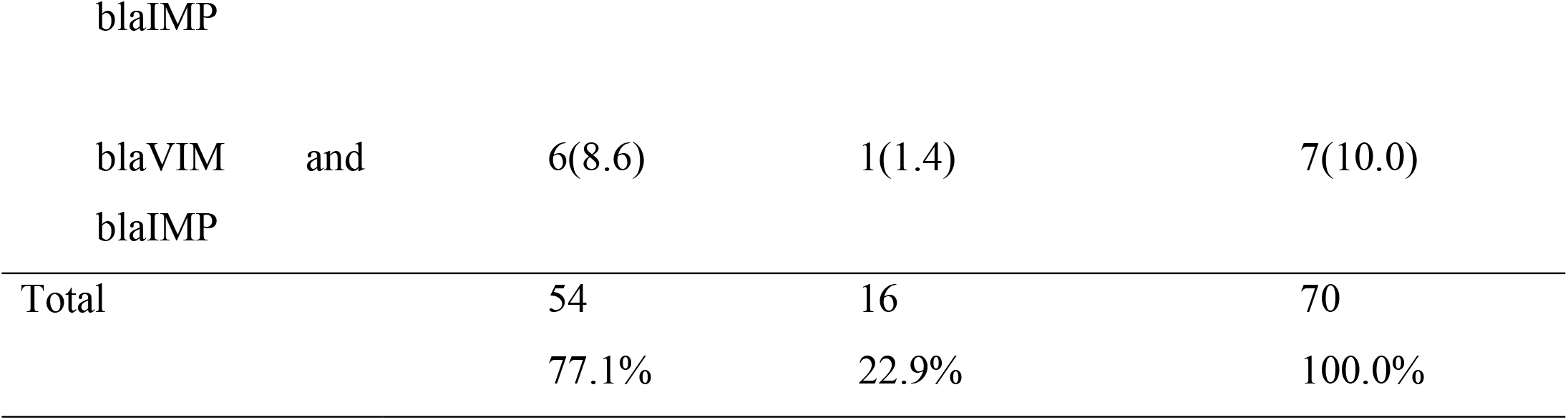
Frequency of four selected carbapenem resistance genes in *C. coli* and *C. jejuni*.

**Fig 2:**
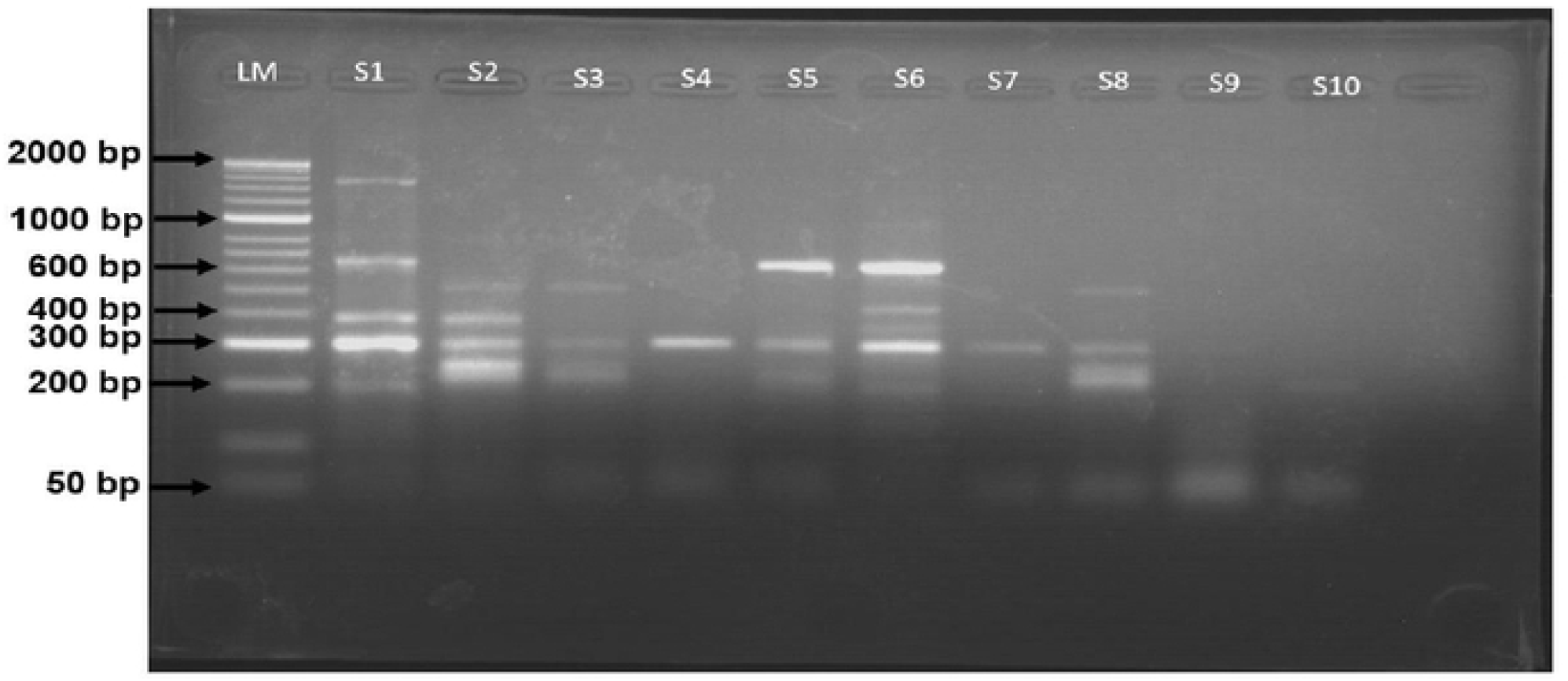
Representative agarose gel for multiplex PCR amplification of selected carbapenem resistance genes (blaVIM, blaIDM-1, blaOXA-48, and blaIMP) in *C. coli* and *C. jejuni* Lane LM: Molecular marker-50bp (Bioline, Alvinston, Canada). Lanes S1-S10 tested samples. Samples S2, S3, S5 and S8 were positive for blaOXA-48; samples S1, S2, and S6 were positive for blaVIM; samples S2, S3 and S8 were positive for blaNDM-1; Samples S1, and S6 were positive for blaIMP. Carbapenem resistance genes; blaNDM-1, blaVIM, blaOXA-48, and blaIMP were detected on a gel by the presence of band sizes of 521bp, 390bp, 238bp, and 587bp, respectively.

## Discussion

Carbapenem resistance is a growing concern in bacterial infections, as these drugs are considered last-resort antibiotics. This study focused on detecting carbapenem resistance genes in *Campylobacter coli* and *Campylobacter jejuni* isolates from children and chicken. Interestingly, all the isolates obtained from diarrheic children and chickens were found to be phenotypically susceptible to Meropenem and Imipenem, which is consistent with the fact that carbapenems are not currently used to treat *Campylobacter* infections in humans and are not licensed for use in animal medicine [18]. This is similar to a study conducted by [19] which reported no resistance to Meropenem and Imipenem in *Campylobacter species* isolated from international travelers. However, it is important to note that with time and increased use of carbapenems, even in animal medicine, resistance is likely to emerge. Although the *Campylobacter* species in this study did not exhibit phenotypic resistance to Meropenem and Imipenem, other pathogens in Uganda have shown resistance to these antibiotics. For example [20] reported prevalence of 33% and 21% in *P. aeruginosa* from patients and the environment respectively while it was 14% and 86% in *A. baumannii* from patients and environment respectively at Mulago hospital. Moreover, [21] reported 22.4% in *Enterobacteriaceae* at Mulago hospital, Kampala. These findings highlight the importance of monitoring resistance patterns in different bacterial species and environments.

The prevalence of carbapenem resistance genes in the *Campylobacter* isolates in this study was 29.79%, which is consistent with previous studies on carbapenem resistance genes in other bacteria in Uganda. For instance, a study on *Enterobacteriaceae* at Mulago Hospital reported a prevalence of 28.6%, while *Acinetobacter baumannii* and *Pseudomonas aeruginosa* showed prevalence rates of 2.7% and 7.4%, respectively [21]. Similar to Enterobacteriaceae, *Campylobacter* species possess mobile genetic elements that can facilitate the horizontal transfer of resistance genes [22]. This emphasizes the need for further research on the mechanisms of gene transfer and integron dynamics to understand the spread and persistence of carbapenem resistance genes in *Campylobacter* in the environment.

The results also revealed that 28.1% (55 out of 196) of *Campylobacte*r isolates from chickens were positive for carbapenem resistance genes, while 38.5% (15 out of 39) of *Campylobacter* isolates from children carried these resistance genes. This prevalence indicates a concerning level of carbapenem resistance genes in both chicken and children’s populations in Kampala city, Uganda. The high prevalence of carbapenem resistance genes in *Campylobacter* isolates from chickens is consistent with previous studies that have identified poultry as a significant reservoir for antimicrobial-resistant bacteria [23]. The poultry industry often employs the widespread use of antibiotics, including critically important carbapenems-like antimicrobials such as cephalosporins, to promote growth and prevent diseases in chickens [24]. Such practices create a favorable environment for the selection and persistence of resistant bacteria in chicken populations.

The higher prevalence of carbapenem resistance genes in Campylobacter isolates from children is particularly concerning. It raises the possibility of release into the environment and transmission of drug resistant *Campylobacter* from human sources to poultry and vice versa possibly through the consumption of contaminated feeds/food and contact with infected chickens. Children, especially those in low-resource settings, are particularly vulnerable to the infection [25], and the presence of carbapenem-resistant *Campylobacter* in this population raises significant public health concerns.

Among the carbapenem resistance genes detected, blaVIM was the most prevalent at 57.1%, which aligns with a previous study that reported blaVIM as the most common gene at 10.1%[21]. Interestingly, all the detected genes (blaVIM, blaNDM-1, blaIMP, and blaOXA-48) were present in C. coli and C. jejuni isolates, despite these isolates being phenotypically susceptible to Meropenem and Imipenem. This suggests that the development of resistance due to the presence of carbapenems in the environment was unlikely as previous studies have indicated that resistance genes can be expressed in the presence of antibiotics in the environment [26]. The high prevalence of blaVIM in this study is suggestive of the role of the accessory genome in evolution of carbapenem resistance in the environment. The blaVIM is characterized as an acquired MBL gene translocated between bacteria via class-1 integrons, in horizontal gene transfer mechanisms mostly involving conjugative plasmids [27] & [22]. This makes it essential to conduct further research on the mechanisms of horizontal gene transfer and integron dynamics. Understanding the molecular pathways of gene transfer will provide insights into the spread and persistence of carbapenem resistance genes in the environment, enabling the development of targeted interventions and control strategies. It is also important to conduct regular monitoring of environmental sources for the presence of carbapenem resistance genes. Regular monitoring of environmental sources, such as wastewater, agricultural runoff, and other potential reservoirs, is also crucial to assess the extent of environmental contamination and identify possible transmission routes.

The study detected a low prevalence of blaOXA-48 at 2.9% among the isolates. This can be attributed to the unique characteristics of OXA-48-like enzymes, which have low or no hydrolytic activity towards carbapenems [28]. Additionally, the identification of OXA-48-type producers can be challenging due to their point mutant analogs with extended-spectrum beta-lactamases (ESBLs) [29]. Hence, estimating the true prevalence rates of OXA-48-like producers is difficult. This study also found co-existence of blaVIM, blaNDM, and blaIMP in *C. coli* and *C. jejuni* isolates at 9(12.9%), indicating that the isolates have the potential for resistance to multiple carbapenem drugs. Although carbapenems have never been licensed for use in livestock animals in any country [30], sporadic cases of carbapenemase-producing bacteria have not only been reported in companion animals but also in livestock, food, and wild animals [31]. Therefore, besides nosocomial bacteria, the food chain and other reservoirs may contribute to the transmission of carbapenem resistance [32]. Strict regulations and discouraging the use of carbapenems in animals are crucial to prevent the emergence and spread of carbapenem resistance genes at the livestock-human interface.

## Conclusions

This study revealed that all the isolates were susceptible to Meropenem and Imipenem. This study also has, for the first time, reported a high prevalence of carbapenem resistance genes in *C. coli* and *C. jejuni* globally at 29.8%. The blaVIM was the most prevalent carbapenem resistance gene.

## Limitation of the study

Samples were collected from limited geographic space, mainly hospital based, and the number was quite small. This impacted the generalizability of the results.

## Recommendations

Further research and analysis to identify and characterize *Campylobacter* species other than *C. coli* and *C. jejuni* should be conducted in such settings as in this study. This investigation should focus on understanding their genetic diversity, resistance gene profiles, and potential involvement in carbapenem resistance. A better understanding of these species will aid in developing targeted interventions and control strategies.

A comprehensive assessment of the current utilization and management practices of carbapenems in both human and animal settings in Uganda should be conducted. This assessment should include surveys, interviews, and data collection from healthcare facilities, veterinary clinics, and farms to determine the extent of carbapenem usage, prescribing patterns, and adherence to guidelines. This information will help identify potential sources of carbapenem resistance and inform future intervention strategies.

A comprehensive surveillance system to monitor the prevalence and trends of carbapenem resistance specifically in *Campylobacter* isolates from diarrheic children and chickens should be established. This surveillance should involve routine testing of isolates for carbapenem resistance genes and phenotypic susceptibility. Timely data collection and analysis will facilitate early detection of emerging resistance patterns and inform targeted interventions.

A robust antibiotic stewardship programs in healthcare and veterinary settings in Uganda should be developed and implemented. These programs should focus on promoting responsible use of carbapenems, ensuring appropriate prescribing practices, and monitoring antimicrobial consumption. Collaboration between human and animal health sectors is crucial to address the potential overlap in antibiotic usage and to harmonize stewardship efforts.

## Data Availability

All relevant data are within the manuscript and its Supporting Information files.

## Authors’ contributions

WO, AN, and TO: conceptualization. WO, TO, AN, JO and FLO: data collection. WO, JO, AN, and TO: formal analysis. OW, TO, and AN: investigations. WO, AN, CK and TO: methodology. WO, AN, TO: discussion. WO, CK, AN, PM, LN, DO, JO, and FLO: writing original draft. WO, JO, AN, LN, and TO: writing final draft. PP, CK, and TO: resources. CK, AN, TO, and PP: supervision. All authors contributed to the article and approved the submitted version.

## Acknowledgment

Our sincere appreciation to the Norway government through NORHED II and the Department of Population Health Sciences, Virginia-Maryland College of Veterinary Medicine for funding this research. A special appreciation also to the staff of Molecular Biology and Food Hygiene Laboratories of COVAB, Makerere University for the technical assistance during the laboratory phase of this study.

## References

[1] F. Prestinaci, P. Pezzotti, and A. Pantosti, “Antimicrobial resistance: a global multifaceted phenomenon,” Pathog. Glob. Health, vol. 109, no. 7, pp. 309–318, 2015.

[2] W. H. Organization, Antimicrobial resistance: global report on surveillance. World Health Organization, 2014.

[3] D. van Duin and Y. Doi, “The global epidemiology of carbapenemase-producing Enterobacteriaceae,” Virulence, vol. 8, no. 4, pp. 460–469, 2017.

[4] M. Kimaanga, “Temporal dynamics of enterobacteriaceae antimicrobial resistance at the human-pig interface in peri-urban Kampala.” Makerere University, 2019.

[5] N. O. Kaakoush, N. Castaño-Rodríguez, H. M. Mitchell, and S. M. Man, “Global epidemiology of Campylobacter infection,” Clin. Microbiol. Rev., vol. 28, no. 3, pp. 687–720, 2015.

[6] C. A. Whitehouse, S. Zhao, and H. Tate, “Antimicrobial resistance in Campylobacter species: mechanisms and genomic epidemiology,” Adv. Appl. Microbiol., vol. 103, pp. 1– 47, 2018.

[7] J. Kim et al., “Comparative analysis of aerotolerance, antibiotic resistance, and virulence gene prevalence in Campylobacter jejuni isolates from retail raw chicken and duck meat in South Korea,” Microorganisms, vol. 7, no. 10, p. 433, 2019.

[8] S. A. Khan, M. A. Imtiaz, M. A. Sayeed, A. H. Shaikat, and M. M. Hassan, “Antimicrobial resistance pattern in domestic animal-wildlife-environmental niche via the food chain to humans with a Bangladesh perspective; a systematic review,” BMC Vet. Res., vol. 16, pp. 1–13, 2020.

[9] H. J. Morrill, J. M. Pogue, K. S. Kaye, and K. L. LaPlante, “Treatment options for carbapenem-resistant Enterobacteriaceae infections,” in Open forum infectious diseases, 2015, vol. 2, no. 2.

[10] M. F. Mushi, S. E. Mshana, C. Imirzalioglu, and F. Bwanga, “Carbapenemase genes among multidrug resistant gram negative clinical isolates from a tertiary hospital in Mwanza, Tanzania,” Biomed Res. Int., vol. 2014, 2014.

[11] L. K. Williams, L. C. Sait, T. A. Cogan, F. Jørgensen, R. Grogono-Thomas, and T. J. Humphrey, “Enrichment culture can bias the isolation of Campylobacter subtypes,” Epidemiol. Infect., vol. 140, no. 7, pp. 1227–1235, 2012.

[12] J. Hudzicki, “Kirby-Bauer disk diffusion susceptibility test protocol,” Am. Soc. Microbiol., vol. 15, pp. 55–63, 2009.

[13] I. I. Lewis and S. James, “Performance standards for antimicrobial susceptibility testing,” (No Title), 2022.

[14] B. da S. Frasao, V. A. Marin, and C. A. Conte-Junior, “Molecular detection, typing, and quantification of Campylobacter spp. in foods of animal origin,” Compr. Rev. Food Sci. Food Saf., vol. 16, no. 4, pp. 721–734, 2017.

[15] D. Linton, A. J. Lawson, R. J. Owen, and J. Stanley, “PCR detection, identification to species level, and fingerprinting of Campylobacter jejuni and Campylobacter coli direct from diarrheic samples,” J. Clin. Microbiol., vol. 35, no. 10, pp. 2568–2572, 1997.

[16] R. Wang, M. F. Slavik, and W. Cao, “A RAPID PCR METHOD FOR DIRECT DETECTION of LOW NUMBERS of CAMPYLOBACTER JEJUNI 1,” J. Rapid Methods Autom. Microbiol., vol. 1, no. 2, pp. 101–108, 1992.

[17] S. A. Khalaf, N. N. Mahmood, and M. A. D. Saleh, “Detection of tetM, armA, blaPER-1 and blaIMP genes in E. coli isolates among the gram negative bacteria that cause urinary tract infections,” in Journal of Physics: Conference Series, 2021, vol. 1999, no. 1, p. 12021.

[18] M. Bassetti, M. Peghin, and D. Pecori, “The management of multidrug-resistant Enterobacteriaceae,” Curr. Opin. Infect. Dis., vol. 29, no. 6, pp. 583–594, 2016.

[19] A. Post et al., “Antibiotic susceptibility profiles among Campylobacter isolates obtained from international travelers between 2007 and 2014,” Eur. J. Clin. Microbiol. Infect. Dis., vol. 36, no. 11, pp. 2101–2107, 2017.

[20] D. P. Kateete, R. Nakanjako, M. Okee, M. L. Joloba, and C. F. Najjuka, “Genotypic diversity among multidrug resistant Pseudomonas aeruginosa and Acinetobacter species at Mulago Hospital in Kampala, Uganda,” BMC Res. Notes, vol. 10, no. 1, pp. 1–10, 2017.

[21] D. Okoche, B. B. Asiimwe, F. A. Katabazi, L. Kato, and C. F. Najjuka, “Prevalence and characterization of carbapenem-resistant Enterobacteriaceae isolated from Mulago National Referral Hospital, Uganda,” PLoS One, vol. 10, no. 8, p. e0135745, 2015.

[22] V. L. Kung, E. A. Ozer, and A. R. Hauser, “The accessory genome of Pseudomonas aeruginosa,” Microbiol. Mol. Biol. Rev., vol. 74, no. 4, pp. 621–641, 2010.

[23] C. M. Schroeder, D. G. White, and J. Meng, “Retail meat and poultry as a reservoir of antimicrobial-resistant Escherichia coli,” Food Microbiol., vol. 21, no. 3, pp. 249–255, 2004.

[24] C. H. Brower et al., “The prevalence of extended-spectrum beta-lactamase-producing multidrug-resistant Escherichia coli in poultry chickens and variation according to farming practices in Punjab, India,” Environ. Health Perspect., vol. 125, no. 7, p. 77015, 2017.

[25] S. Budge et al., “Risk factors and transmission pathways associated with infant Campylobacter spp. prevalence and malnutrition: A formative study in rural Ethiopia,” PLoS One, vol. 15, no. 5, p. e0232541, 2020.

[26] H. Knothe, P. Shah, V. Krcmery, M. Antal, and S. Mitsuhashi, “Transferable resistance to cefotaxime, cefoxitin, cefamandole and cefuroxime in clinical isolates of Klebsiella pneumoniae and Serratia marcescens,” Infection, vol. 11, no. 6, pp. 315–317, 1983.

[27] G. J. Da Silva and S. Domingues, “Insights on the horizontal gene transfer of carbapenemase determinants in the opportunistic pathogen Acinetobacter baumannii,” Microorganisms, vol. 4, no. 3, p. 29, 2016.

[28] M. Rivera-Izquierdo et al., “OXA-48 carbapenemase-producing enterobacterales in Spanish hospitals: An updated comprehensive review on a rising antimicrobial resistance,” Antibiotics, vol. 10, no. 1, p. 89, 2021.

[29] F. S. Codjoe and E. S. Donkor, “Carbapenem resistance: a review,” Med. Sci., vol. 6, no. 1, p. 1, 2017.

[30] J. Fernández, B. Guerra, and M. R. Rodicio, “Resistance to carbapenems in non-typhoidal Salmonella enterica serovars from humans, animals and food,” Vet. Sci., vol. 5, no. 2, p. 40, 2018.

[31] N. Woodford, D. W. Wareham, B. Guerra, and C. Teale, “Carbapenemase-producing Enterobacteriaceae and non-Enterobacteriaceae from animals and the environment: an emerging public health risk of our own making?,” J. Antimicrob. Chemother., vol. 69, no. 2, pp. 287–291, 2014.

[32] I. Kühn et al., “Epidemiology and ecology of enterococci, with special reference to antibiotic resistant strains, in animals, humans and the environment: example of an ongoing project within the European research programme,” Int. J. Antimicrob. Agents, vol. 14, no. 4, pp. 337–342, 2000.

